# Impact of pediatric influenza vaccination on antibiotic resistance in England and Wales

**DOI:** 10.1101/19003657

**Authors:** Chungman Chae, Nicholas G. Davies, Mark Jit, Katherine E. Atkins

## Abstract

Vaccines against viral infections have been proposed to reduce antibiotic prescribing and thereby help control resistant bacterial infections. However, by combining published data sources, we predict that pediatric live attenuated influenza vaccination in England and Wales will not have a major impact upon antibiotic consumption or health burdens of resistance.

---

Antibiotic use drives the spread of antibiotic resistance. A substantial proportion of antibiotic prescriptions are unnecessary because they are written to treat conditions that are either self-limiting or non-bacterial in etiology (*1*). Since influenza is often treated inappropriately with antibiotics, expanding access to influenza vaccines has been proposed as a means of reducing unnecessary prescribing and preventing resistant infections (*2*).

In 2013, England and Wales began rolling out the live attenuated influenza vaccine (LAIV) for 2–16-year-old children (*3*). Here, we estimate the potential impact on antibiotic prescribing and antibiotic resistance.

## Methods

We assume that some influenza cases lead to general practitioner (GP) consultations and some GP consultations lead to antibiotic prescriptions. We focus on community antibiotic use as the driver of resistance, as hospitalizations for influenza are rare relative to GP consultations (*4*).

### Influenza-attributable consultations

For our base-case influenza-attributable consultation rate, we use previous estimates from a time-series statistical attribution analysis over the 1995–2009 UK flu seasons (*5*), yielding an average of 14.7 influenza-attributable GP consultations per 1000 person-years (Table 1). For our uncertainty analysis, we use a lower estimate of 11.8 per 1000, derived from a longitudinal study in England over 2006–2011 (*6*), and a higher estimate of 21.4 per 1000, derived from a time-series statistical analysis for England and Wales over 2000–2008 (*4*).

**Table 1.** Projected impact of pediatric LAIV on antibiotic prescription rates, England and Wales*

| Age group | Influenza-attributed consultation rate† | Prescriptions per consultation | Direct prescribing rate reduction, unmatched‡ | Direct prescribing rate reduction, matched‡ | LAIV impact§ | Overall prescribing rate reduction¶ |
| --- | --- | --- | --- | --- | --- | --- |
| 0–6m | 29.7 (23.7 – 35.9) | 0.597 (0.474 – 0.719) | — | — | 0.574 (0.501 – 0.651) | 10.2 (7.03 – 13.5) |
| 6m–4y | 29.7 (23.7 – 35.9) | 0.597 (0.474 – 0.719) | 7.46 (5.31 – 9.64) | 12.4 (8.85 – 16.1) | 0.663 (0.618 – 0.714) | 11.8 (8.31 – 15.4) |
| 5–14y | 22.1 (17.6 – 26.7) | 0.588 (0.466 – 0.708) | 5.46 (3.89 – 7.06) | 9.11 (6.48 – 11.8) | 0.754 (0.709 – 0.794) | 9.81 (6.97 – 12.8) |
| 15–44y | 12.8 (10.2 – 15.4) | 0.676 (0.536 – 0.814) | 3.64 (2.59 – 4.70) | 6.06 (4.31 – 7.83) | 0.446 (0.394 – 0.502) | 3.86 (2.66 – 5.09) |
| 45–64y | 12.4 (9.84 – 14.9) | 0.805 (0.639 – 0.970) | — | — | 0.423 (0.374 – 0.484) | 4.22 (2.90 – 5.58) |
| 65y+ | 12.2 (9.67 – 14.7) | 0.857 (0.680 – 1.03) | — | — | 0.477 (0.397 – 0.561) | 4.97 (3.34 – 6.68) |
| Overall | 14.7 (11.7 – 17.7) | 0.726 (0.576 – 0.875) | 5.80 (4.13 – 7.49) | 9.86 (7.01 – 12.9) | 0.494 (0.446 – 0.549) | 5.32 (3.74 – 7.00) |

\*All estimates reported as mean (95% highest density interval).
†Per 1000 people per year in England and Wales.
‡Reduction in antibiotic prescriptions among vaccinees per 1000 vaccine recipients, not accounting for herd immunity, separately for unmatched and matched seasons.
§Reduction in influenza cases assuming a 50% uptake among 2–16-year-olds, accounting for herd immunity.
¶Per 1000 people per year in England and Wales, accounting for herd immunity.

### Prescriptions per consultation

For our base-case analysis we use a previous UK average estimate of 726 antibiotic prescriptions for every 1000 influenza-attributable GP consultations (*5*). For our uncertainty analysis, we use a lower estimate of 313 per 1000, based on electronic health records of prescriptions within 30 days of a consultation for influenza-like illness (ILI) or acute cough in England over 2013–2015 (*7*).

### Vaccine impact

In our base-case analysis, we assume that LAIV prevents 49% of symptomatic influenza cases on average, from a previously published mathematical model of pediatric LAIV in England and Wales which assumes 50% uptake and either 70% (matched years) or 42% (unmatched years) efficacy among 2–16-year-olds (*3*). This reduction is consistent with a pilot study comparing consultation rates in treatment versus control areas before and after LAIV rollout (*8*). For our uncertainty analysis, we use lower and upper estimates of 32% and 63% fewer influenza cases from the same model, assuming an uptake of 30% and 70%, respectively.

### Link between antibiotics and resistance

We use linear regression to predict the per-country health burden of each resistant strain as a function of the country’s rate of primary-care antibiotic consumption, using previously published data across EU/EEA countries in 2015 for 16 resistant bacterial strains (*9*). The regression slope predicts the impact of reducing antibiotic consumption by a defined amount.

### Economic cost of resistance

We adopt a published cost estimate of $1415 per resistant infection (2016 USD) (*10*), adjusted for inflation and health care purchasing power parity for the UK to £520 (2015 GBP).

### Further details

We use Monte Carlo sampling to explore uncertainty across estimates for consultation rate, prescribing rate and LAIV impact, weighting age groups using demographics for England and Wales in 2015. Analysis code and data are available at github.com/nicholasdavies/laiv_amr_ew. Additional details are in the Appendix.

## Results

We find that LAIV administered to 2–16-year-olds has the potential to reduce antibiotic consumption by 5.3 (95% highest density interval: 3.7–7.0) prescriptions per 1000 person-years (Table 1) across the population of England and Wales. This is equivalent to 0.8% of the total antibiotic dispensation rate in English primary care in 2015. For comparison with secular trends, this rate has fallen by 2.5% each year from 2012 to 2018 (Appendix Figure 1). Focusing on vaccine recipients only, we estimate that the direct effectiveness of LAIV on antibiotic consumption is 5.8 (4.1 – 7.5) fewer prescriptions per thousand person-years in unmatched years and 9.9 (7.0 – 13) in matched years.

**Figure 1.**
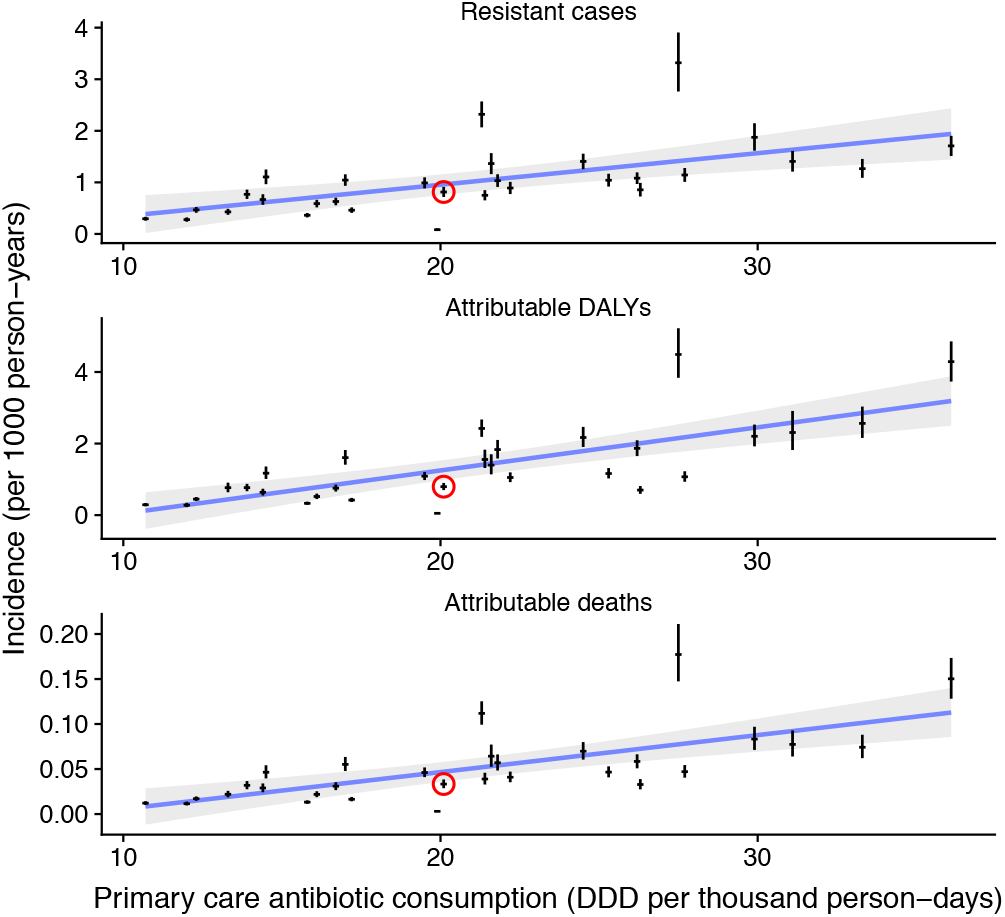
Estimated health burdens of antimicrobial resistance plotted against the overall antibiotic consumption in primary care for 30 European countries, 2015. The United Kingdom is circled.

Although 0.8% is a small decrease in antibiotic use, there may be an appreciable impact on the cost-effectiveness of pediatric LAIV if the health burdens of resistance are substantial enough (Figure 1). We estimate that LAIV has the potential to reduce resistance-attributable disability-adjusted life years (DALYs) by 642, cases by 432 and deaths by 22 per year in England and Wales (Table 2), with averted DALYs spread relatively evenly across the 7 causative pathogen species analyzed (Figure 2A). We estimate a yearly cost saving of £224K for averted resistant infections. When compared with the projected incremental cost (program cost minus health care saving) of pediatric LAIV at £63.6M, and its projected impact of saving 27,475 quality-adjusted life years (QALYs) and averting 799 deaths yearly (*3*), accounting for resistance will not substantially increase the cost-effectiveness of pediatric LAIV in this setting. Our uncertainty analysis (Figure 2B) identifies the consultation rate as having the greatest influence over the impact of LAIV on health burdens of resistance.

**Table 2.** Projected impact of pediatric LAIV on health burdens of antibiotic resistance, England and Wales*

|  | Estimated 2015 burden | Projected reduction in burden from LAIV |
| --- | --- | --- |
| DALYs | 46039 | 432 (303 – 566) |
| Cases | 47080 | 642 (450 – 842) |
| Deaths | 1930 | 22 (16 – 29) |

\*Reductions reported as mean (95% highest density interval).

**Figure 2.**
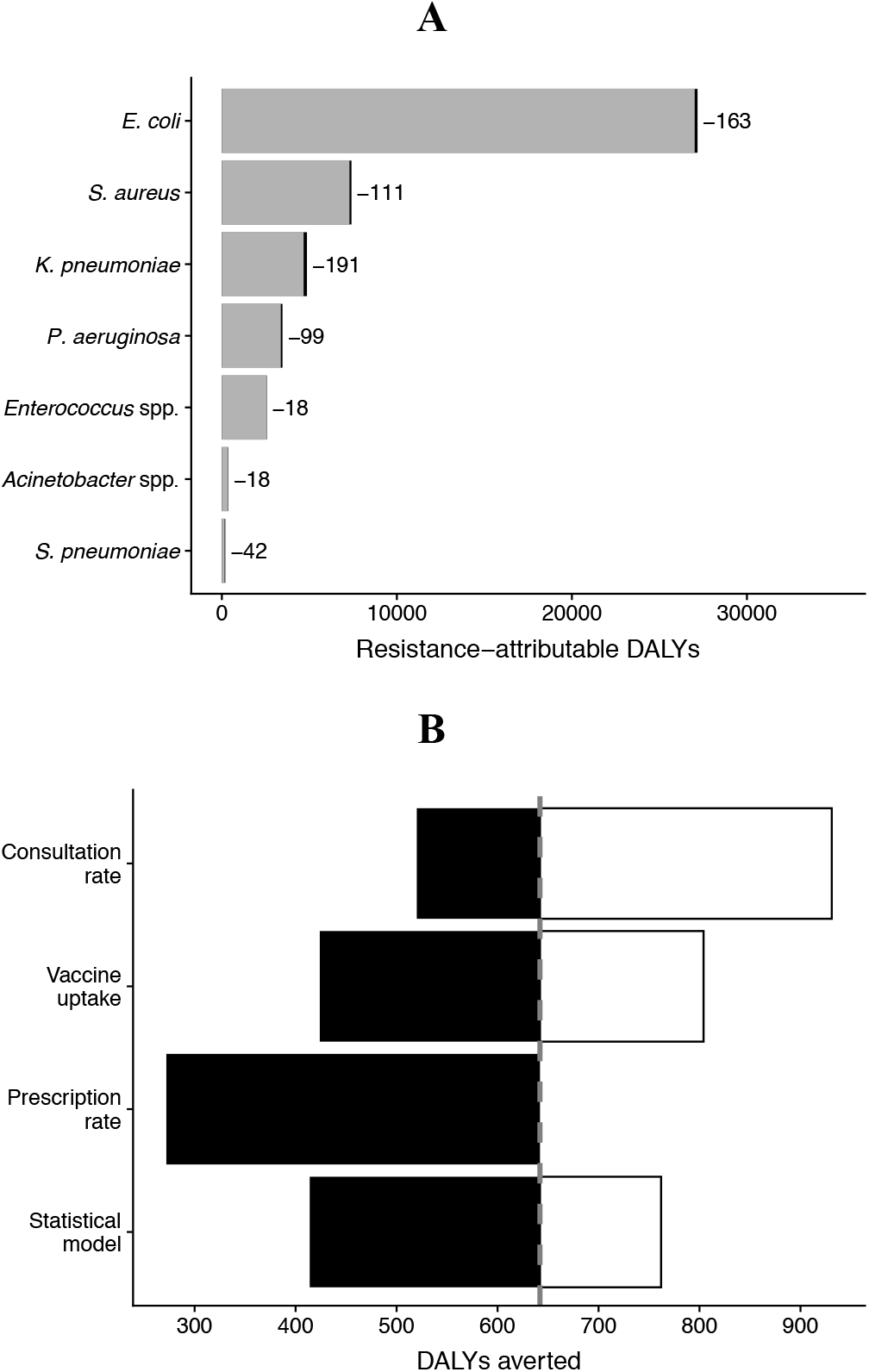
A) Estimated DALYs averted by pediatric LAIV in England and Wales, stratified by causative pathogen; the entire width of the bar is the current burden, with potential reductions highlighted in black and reported next to each bar. B) One-way uncertainty analysis, showing the impact on DALYs averted of alternative assumptions concerning the rate of influenza-attributable GP consultations, the pediatric uptake of LAIV, the rate of antibiotic prescribing per GP consultation, and how the impact of prescribing on health burdens is attributed (see Methods and Appendix for more details).

## Discussion

Our estimates for the foreseeable reduction in antibiotic prescribing from the LAIV program in England and Wales may seem surprisingly low, given that sore throat, cough, and sinusitis together account for 53% of all inappropriate prescribing, which in turn comprises at least 9–23% of all prescribing in England (*1*). However, many viral and bacterial pathogens cause these symptoms. By one estimate, influenza causes only 11% of GP consultations for acute respiratory illness in England (*4*), so it may be optimistic to expect influenza vaccination to substantially reduce antibiotic use in the UK.

Our base-case estimate of 726 antibiotic prescriptions per 1000 influenza-attributable consultations is more than double what electronic health records suggest (Methods). One explanation is that this estimate, derived from statistical attribution of antibiotic prescriptions to influenza circulation over 1995–2009 (*5*), feasibly includes prescribing for infections secondary to influenza infection, such as otitis media, sinusitis and pneumonia. Also, antibiotic use in England has declined since this time—by 22% from 1998 to 2016 (*11*). Accordingly, our base-case results should be interpreted as the maximum potential reduction by LAIV of antibiotic use. Conversely, in the FluWatch study only 8% of consultations for ILI resulted in influenza or ILI being medically recorded (*6*), and so electronic health records may not reliably reflect prescribing rates for influenza.

In randomized trials, the direct effect of influenza vaccines on vaccinated children has ranged from a 44% reduction in antibiotic prescriptions (Italy) to a 6% increase (United States), both over the 4-month period following vaccination (*12*). Estimates of the impact over entire populations (all ages, vaccinated and unvaccinated) range from 11.3 fewer prescriptions per 1000 person-years in Ontario (*13*) to 3.9 fewer in South Africa and Senegal (*14*). This variation can be ascribed to differences in vaccine efficacy and coverage, influenza circulation, existing patterns of antibiotic use, and methodology; estimates of vaccine impact on antibiotic consumption may not be generalizable across settings.

Consistent with our results, UK-specific empirical estimates suggest little or no effect of LAIV on prescribing. A self-controlled case series study found that 2–4-year-old LAIV recipients in the UK took 13.5% fewer amoxicillin courses in the 6 months following vaccination (*15*), while an LAIV pilot study detected no difference in prescribing rates for respiratory tract infections between treatment groups (*16*). No single vaccine is likely to substantially reduce inappropriate antibiotic use in the UK.

## Data Availability

Analysis code and data are available at github.com/nicholasdavies/laiv_amr_ew.

## Acknowledgements

We thank Edwin van Leeuwen for providing results from the mathematical model of influenza transmission and vaccination, Diamantis Plachouras for correspondence, and David R. M. Smith, Edwin van Leeuwen, and Marc Baguelin for discussion.

## Transparency declarations

Nothing to declare.

## Appendix

### 1. Influenza-attributable GP visits

#### 1.1 Base-case estimate

From Fleming et al. (*1*) Table 2, consultations for respiratory disease broadly defined attributable to either influenza A or B. Age groups reported by Fleming et al. differ from those used in this study, so we adapt estimates of Fleming et al. by assuming that reported rates are constant within an age group, and that half of children under 12 months old are under 6 months old.

Fleming et al. do not directly report confidence intervals in measured rates (instead, variation between flu seasons is reported), so we assume that uncertainty in the influenza-attributable GP consultation rates follows a normal distribution. We assume that the standard deviation of any consultation rate derived from this source is always *S* times the mean rate, where *S* is estimated from Fig. 2 of Fleming et al. (*1*) by assuming that the width of the 95% confidence intervals on this figure are equivalent to 1.96 times the standard deviation of an associated normal distribution, and that the standard deviation of influenza-attributable GP visits is 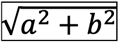 where *a* is the standard deviation of influenza A-attributable consultations and *b* is the standard-deviation of influenza B-attributable consultations. *S* is then the mean relative standard deviation, calculated in this manner, across all study years.

#### 1.2 Low estimate

Rates of PCR confirmed influenza are estimated from Hayward et al. (*2*), Tables S2 and S3, taking the mean over the five winter flu seasons reported (i.e. excluding the Summer 2009 pandemic flu period), assuming that the reported rates of PCR confirmed influenza in the form B (A – C) represent a triangular distribution with B as the peak (mode) and A – C as the 95% highest density interval (using a triangular distribution rather than a normal distribution allows us to account for skew). Then, the probability of a GP visit given PCR-confirmed illness is taken from Table S6 of the same source. We correct for low numbers by assuming a “base proportion” of 12/82 as the measured proportion of PCR confirmable influenza episodes resulting in a GP visit, which comes from the overall number of reported GP visits for 5–64-year-olds with PCR-confirmed influenza. To account for uncertainty in measurement, we draw the “base rate” of GP consultation given PCR-confirmable influenza for 5–64-year-olds from a beta distribution with parameters *α* = 12 + 1,*β* = 82−12 + 1 (i.e. assuming a uniform prior); to account for the observation that this rate is higher in young children and the elderly (Table S6), we add 0.12 to this rate for under-5s and over-65s. The annual influenza-attributable rate of GP consultation for a given age group is then the product of the PCR-confirmable influenza incidence and the rate of GP consultation given PCR-confirmable influenza.

#### 1.3 High estimate

These are taken from Cromer et al. (*3*), Table 4, assuming that reported 95% confidence intervals represent 1.96 times the standard deviation of a normal distribution.

### 2. Rate of antibiotic prescribing given an influenza-attributable GP consultation

#### 2.1 Base-case estimate

From Fleming et al. (*1*) Table 2, by dividing the rate of antibiotic prescribing by the rate of influenza-attributable GP consultations, assuming a normal distribution for the final rate with the same relative standard deviation derived in 1.1 above.

#### 2.2 Low estimate

From Pouwels et al. (*4*) Table 3, which reports that 48% of consultations for acute cough and 29% of consultations for influenza-like illness result in a systemic antibiotic prescription within 30 days. We assume that 88.1% of influenza-attributable consultations are for ILI (hence having a 29% prescription rate) and the rest are for acute respiratory infection without fever (*2*) (hence having a 48% prescription rate), which yields an overall (crude) prescribing rate of 31.3%.

To calculate age-stratified values, we assume prescribing for under-5s is about 20% less, and for over-45s is about 20% more, than prescribing in 5–44-year-olds, consistent with the results of Fleming et al. (*1*), Meier et al. (*5*), and Pitman et al. (*6*). That is, we draw a value *d* from a normal distribution with mean 0.2 and standard deviation 0.05, and assume that the relative prescribing rate for under-5s is (1 – *d*) times the rate for 5–44-year-olds, while the relative prescribing rate for over-45s is (1 + *d*) times the rate for 5–44-year-olds.

### 3. Impact of LAIV on rates of GP consultation

We use fitted models from Baguelin et al. (*7*) projecting the impact of LAIV on influenza cases in different age groups, assuming either a 50% uptake (base-case estimate), 30% uptake (low estimate), or 70% uptake (high estimate).

### 4. Prediction of prescription rate impact on resistance burdens

#### 4.1 Defined daily doses per prescribed antibiotic course

We assume that each prescription comprises 7 defined daily doses (DDD), as 7 days is the typical duration of antibiotic treatment for upper respiratory tract infections (*8*).

#### 4.2 Main scenario

We use total primary care antibiotic consumption (ATC code J01C) for European countries for 2015 from the ECDC (*9*) as the predictor variable, and per-country median health burden (DALYs, cases, or deaths) attributed to each of 16 strains analyzed by Cassini et al. (*10*) as the outcome variable, in a series of country-level linear regressions from which we separately predict the impact of reducing overall prescribing by a defined amount.

#### 4.3 Alternative scenario 1

Rather than using the overall antibiotic consumption, we also built a separate series of models where we used as predictors each country’s consumption of tetracyclines (J01AA), extended spectrum penicillins (J01CA), beta-lactamase sensitive penicillins (J01CE), and macrolides (J01FA), as these four classes comprise the majority of antibiotics prescribed for sore throat and cough (*11*). We assume that for a given reduction in the overall prescription rate *x*, there is a reduction 0.0620*x* in tetracycline prescribing, 0.4752*x* in extended spectrum penicillin prescribing, 0.2793*x* in beta-lactamase sensitive penicillin prescribing, and 0.1835*x* in macrolide prescribing. This predicted a smaller impact upon resistance than the main scenario (4.1) and comprises the “low-effect” statistical model for the uncertainty analysis (Fig. 2B, main text).

#### 4.4 Alternative scenario 2

We follow the same procedure as in 4.2, but if any predictor variable is negatively correlated with a resistance related health burden (i.e. the best fitting linear model suggests that decreasing use of that antibiotic would increase resistance), we remove it from the linear regression and rerun the model, continuing this process until all predictors are positively associated with the outcome variable. If more than one variable has a negative association in a given round, all are removed for the next round. This predicted a larger impact upon resistance than the main scenario (4.1) and comprises the “high-effect” statistical model for the uncertainty analysis (Fig. 2B, main text).

### 5. Economic calculations

To convert between US and UK health care expenditures, we use hospital-service price level indices for health care purchasing power parity published by the OECD (*12*) (see their Fig. 1).

### 6. Secular trends in antibiotic prescribing rates

Data from NHS Digital show that community antibiotic use in England has decreased by approximately 2.5% per year from 2012 to 2018 (Appendix Figure 1).

**Appendix Figure 1.**
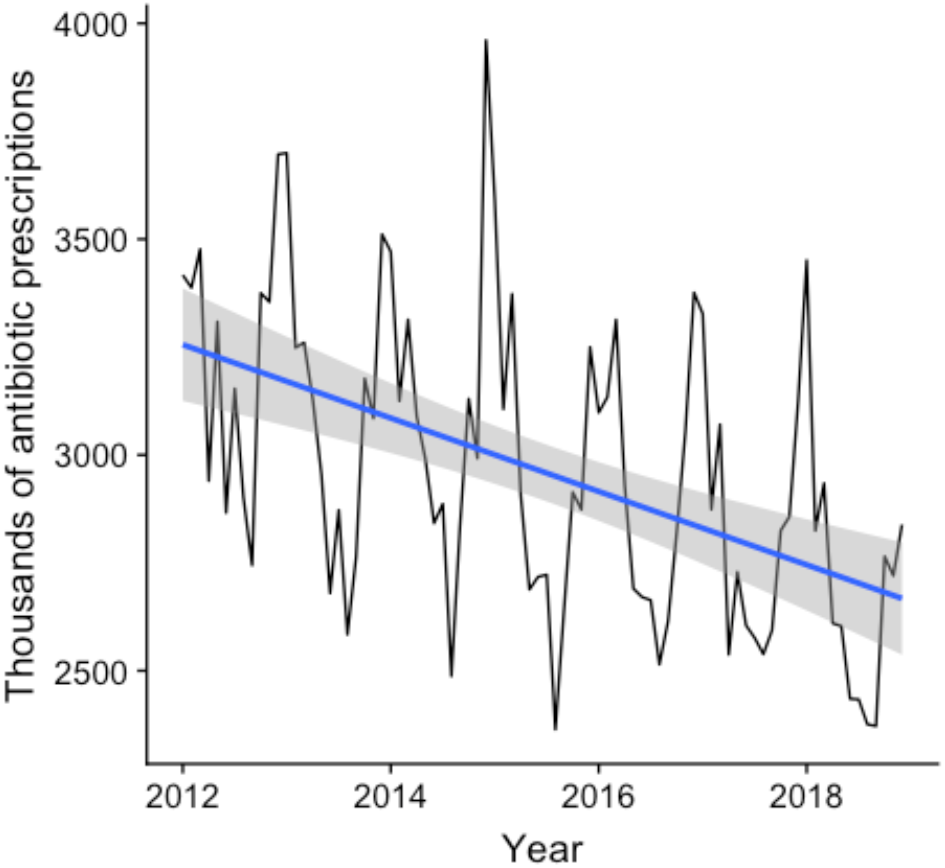
Antibiotic use in England has fallen by approximately 2.5% each year from 2012 to 2018.

